# Trend of new cases of Human Immunodeficiency Virus infections in two health facilities in the Northern Cameroon between 2021-2022

**DOI:** 10.1101/2024.04.10.24305613

**Authors:** Patrice Djataou, Marceline Ngounoue Djuidje, Georges Nguefack-Tsague, Jean de Dieu Anoubissi, Joel Kadji Kameni, Aline Tiga, Elise Elong, Moussa Djaouda, Alexis Ndjolo, Céline Nguefeu Nkenfou

## Abstract

HIV/AIDS continues to be a global public health problem. Studies of the incidence and prevalence of HIV and other sexually transmitted infections (STIs) that may contribute to or aggravate its acquisition remain an effective means of prevention. In recent years, terrorist groups have established themselves in the northern regions of Cameroon. This insecurity has led to a large influx of refugees with no information about their HIV and STI status. Given this above mentioned situation, this study aimed to assess the incidence and prevalence of HIV and STI and their associated risk factors in order to adjust strategies to monitor the epidemic. A cohort of 684 consenting participants from the North and Far North were enrolled in the study in 2021 and followed up in 2022 to measure the incidence and prevalence of HIV and to assess some associated risk factors. Each participant was administered a pretested questionnaire to collect sociodemographic variables and risk behaviors. Anti-HIV Ab, HBsAg (Hepatitis B Surface Antigen), TPHA (*Treponema Pallidum* Hemagglutination Assay) tests were performed. The data were compiled using EPI Info 7.5.2 for epidemiological analyses. The association between co-infections of HIV, Hepatitis, and syphilis and HIV incidence was evaluated using the Chi-2 test. The HIV incidence and overall prevalence were 1.63% (163/10,000 population) and 3.8%, respectively. The HIV incidence increased from 0.27% in 2017 (DHS) to 1.63% in the North and Far North regions as found in our study. The incidences of syphilis and hepatitis B were 1.03% and 4.56%, respectively. Factors associated with HIV acquisition included religion (Muslims being more infected, P<0.03), unprotected sex with a new partner (P<0.007), having a sex worker as a partner (P<0.0001), and co-infection with syphilis and hepatitis B (P<0.05). The findings also link increased HIV incidence to insecurity and population displacement. In HIV prevention strategies, it is important to consider the security and political stability context as well as HIV-associated infections such as hepatitis B and syphilis.

## INTRODUCTION

In 2020, there were still 1.3 million new HIV infections despite considerable efforts to reduce the trend. The total number of people living with HIV (PLHIV) worldwide was 37.7 million, with 20.6 million in Africa [1]. Currently, there is no vaccine or curative treatment for HIV/AIDS. Control of this pandemic still relies on prevention and surveillance. According to UNAIDS’ 2019 95-95-95 strategic plan projected in 2025[2], 95% of people living with HIV should know their HIV status, 95% of all HIV-infected people tested should receive sustained antiretroviral therapy, and 95% of people receiving antiretroviral therapy should have sustained viral load suppression. The study and surveillance of risk factors of incidence, prevalence, and co-infections that facilitate the acquisition of HIV is one effective means of prevention and surveillance that provides information on the epidemiological evolution of HIV/AIDS.

Cameroon is one of the most affected countries in Sub-Saharan Africa with generalized epidemics [3]. Although overall adult HIV prevalence continues to decrease, moving from 5.4% in 2004[4] to 4.3% in 2011[5], 3.4% in 2017[6] and recently 2.7% in 2018[7], continuous monitoring is advised. From data collected, the prevalence is not uniformly distributed across regions. This is related to socio-cultural parameters such as religion, occupation, sexual behavior, neighborhood, traditions or poverty, political security and the associated influx of refugees, or the strength of the health system [8–10]. According to a World Bank report, 56% of poverty in Cameroon occurs in the northern regions alone, where Islam is one of the main religions. HIV prevalence is lowest in the North and Far North regions of Cameroon (1.6% and 1.5%, respectively). In recent years, the terrorist group Boko Haram has established itself in the Far North region of Cameroon. These security issues have led to a significant influx of refugees into the northern part of the country without information on their HIV and STI status. With the massive influx of refugees, this study aimed to assess the incidence and prevalence of HIV and STIs and their associated risk factors in order to adapt the monitoring of the epidemic [11].

## METHODOLOGY

### Study design

We chose to work in the two most accessible hospitals in the North and Far North regions of Cameroon.

The study was conducted at the main entry point of the regional hospitals of Garoua and Maroua, located in the North and Far North regions of Cameroon, respectively. Enrolled participants were tested in the first year and re-tested in the second year, 12 months apart, i.e. four HIV serologic windows (June 2021-May 2022). This study was approved by the National Ethics Committee of Cameroon. The minimum sample size was calculated with a margin of error of 5% according to the following formula [12].

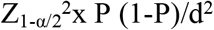

P: Prevalence of HIV infection in north and far north is under 5%

Zα/2: Value of the fractile of the normal distribution at 5%=1.96;

d: Degree of precision = 2%; the calculated size is N=616 participants.

Taking into account a loss to follow-up of 10% (68 participants), the minimum sample size was 684 participants.

The study included consented individuals aged 18-70 years of both sexes who had lived in the North and Far North regions for at least 5 years and who came to the hospital for work, medical consultations or as visitors.

Recruitment was conducted in two phases. Phase 1 (Year 1) ran from June 15, 2021 to August 13, 2021, and Phase 2 (Year 2) ran from May 30, 2022 to July 15, 2022. During the first year of the study, 684 participants were enrolled and 5 mL of blood was drawn for the following tests HIV1/2; HBsAg and TPHA (*Treponema pallidium* heamagglutination assay). These STIs are known to exacerbate HIV infection or facilitate its acquisition.

For HIV1/2, the rapid diagnostic test was performed using Determine Alere HIV 1/2 Strips (Abbott Diagnostics Medical Co., Ltd. 357 Matsuhidai, Matsudo-shi Chiba, 270-2214, Japan +81473115750) and confirmed by KHB (Shanghai Kehua Bio-engineering Co., Ltd. www.skhb.com) cassettes according to the manufacturer’s instructions. HBsAg test was performed using Fortress Diagnostic’s HBsAg rapid screening tests and confirmed using HBsAg CYPRESS diagnostics ELISA kits (Cypress Diagnostics: Langdorpsesteenweg 160.3201 Langdrop, Belgium www.diagnostics.be Tel: +3215676768) according to the manufacturer’s instructions. Syphilis test (TPHA) was performed by SD BIOLINE Syphilis tests and confirmed by ELISA Bio-Rad (Bio-Rad 3, boulevard Raymond Poincaré, 92430 Marnes-la-coquette-France. Tel: +33(0)147956000, www.bio-rad.com) according to the manufacturer’s procedures.

During the second year of the study, of the 684 participants enrolled in the first year, 676 were retested, 06 participants were unable to attend the study site, and the remaining 02 participants were deceased. Participants who tested positive in the first year were not retested. Sociodemographic data were obtained by administering a questionnaire. The chi-squared test was used to compare the prevalences obtained with those known. New cases of infection were also compared with the national incidence, and sociocultural risk factors were analyzed. P<0.05 values were considered statistically significant.

### Ethical considerations

This work was approved by the hospitals and the National Ethics Committee for Human Health in Cameroon (N°2021/06/85/CE/CNRESH/SP). Recruited participants agreed to participate in the study and provided a written informed consent form.

## RESULTS

### Sociodemographic characteristics of the study population

A total of 684 participants were randomly enrolled, regardless of gender, and the mean age was 31.4 ± 2.4 years with a standard deviation of 12 ± 1.5 years. The most represented age group was that of 18 to 35 years 62.33%). Females represented 47.37% (324/684) of our study population compared to 52.6% (360/684) of males. Of these 684 participants, the mean age at first sexual intercourse was (19.66±4.6) years with a standard deviation of (3.7±0.8) years, with a minimum of 12 years and a maximum of 32 years. 15.06% of the participants admitted to having had a sexually transmitted infection such as syphilis (n=41), gonorrhea (n=37) and chlamydia (n=12). These data are shown in Table I.

**Table I:** Socio-demographic and clinical characteristics of the study population.

|  | Men | Women |
| --- | --- | --- |
| **Participants** | 360(52.6%) | 324(47.37%) |
| **Mean Age** | 30.1 ± 2.1 | 32.03 ± 3.3 |
| **Religion** |  |  |
| • Muslim | 129 | 97 |
| • Christian | 231 | 227 |
| **Education level** |  |  |
| • University | 71 | 52 |
| • Secondary | 239 | 128 |
| • Primary | 43 | 18 |
| • illiterate/coranic | 7 | 126 |
| **Matrimonial status** |  |  |
| • Married | 117 | 142 |
| • Single | 156 | 168 |
| • Divorced/widow | 87 | 14 |

Note: school enrolment in northern cameroon is on the rise. However, young girls are still the most marginalized.

### Incidence and prevalence of HIV, Hepatitis B and syphilis in the study population

The prevalence of HIV, syphilis, and hepatitis B infections by sex in the first year of this study were 1.90%, 4.09%, and 0.146%, respectively.

New cases of HIV, hepatitis B, and syphilis infections were 1.63%, 4.72%, and 01.03%, respectively (Table II). Compared with the incidence of HIV in Cameroon, the new infections are statistically significant (P<0.001).

**Tableau II:** Prevalence and incidence of HIV, Hepatitis B, and syphilis in the study population

|  | HIV | Hepatite B | Syphilis |
| --- | --- | --- | --- |
| **Prevalence** | 02.63% | 06.28% | 01.16% |
| **Incidence** | 01.63% | 4.72% | 1.02% |

Note: New cases of HIV infection detected were enrolled in the Region's immediate test and treat program. Cases of new syphilis infections were immediately managed by the medical profession. Hepatitis B cases were referred to the pasteur center for follow-up.

The global incidence of HIV-hepatitis B and HIV-syphilis co-infection was 0.29% and 0.15%, respectively. The prevalence of HIV-syphilis and HIV-hepatitis co-infection in the first year of the study was zero.

### Risk Factors associated to HIV infection

From table III below, religion (Muslim P<0.03), practice of unprotected sex with a new partner (P<0.007), and sexual relationship with sex workers (P<0.0001) were the socio-behavioural factors associated with HIV acquisition.

**Tableau III:** Risk factors associated to HIV infection.

| Variables | HIV- (%) | HIV+ (%) | P-value |
| --- | --- | --- | --- |
| **Age at 1<sup>st</sup> sexual intercourse** |  |  |  |
| ≤18 years (%) | 275 (97.87) | 6 (2.13) | 0.5 |
| >18years (%) | 343 (98.56) | 5 (1.44) |  |
| Never (%) | 44(100) | 00 (0.00) |  |
| **Condom use at 1<sup>st</sup> sexual intercourse** |  |  |  |
| Yes (%) | 352 (98.87) | 04 (1.13) | 0.59 |
| No (%) | 267 (97.45) | 07 (2.55) |  |
| Never had sex (%) | 46 (100) | 00 (0.00) |  |
| **Number of sexual partners** |  |  |  |
| None (%) | 36 (100) | 00 (00) | 0.1 |
| 1 partner (%) | 469 (96.13) | 06 (3.87) |  |
| 2 partners (%) | 114 (97.39) | 03 (2.61) |  |
| ≥ 3 partners (%) | 46 (95.83) | 02 (4.17) |  |
| **Knowledge of the HIV status of the partner(s)** |  |  |  |
| Yes (%) | 351 (71.64) | 08 (28.36) | 0.39 |
| No (%) | 296 (68.49) | 03 (31.51) |  |
| No partner (%) | 18 (100) | 0 |  |
| **Religion** |  |  |  |
| Muslims (%) | 226 (96.58) | 08 (3.42) | **0.027** |
| Christians (%) | 437 (99.32) | 03 (0.68) |  |
| Other (%) | 02 (100) | 00 (00) |  |
| **Past STI** |  |  |  |
| Yes | 99 (98,02) | 02 (1.98) | 0.76 |
| No | 566 (98,43) | 09 (1,57) |  |
| **Unprotected sex with a new partner** |  |  |  |
| Yes (%) | 101 (95.28) | 05 (4.72) | **0.006** |
| No (%) | 564 (98.95) | 06 (1.05) |  |
| **Sexual relationship with sex worker** |  |  |  |
| Yes (%) | 95 (91,35) | 09 (8,65) | **0.0001** |
| No (%) | 256 (98,08) | 05 (1,92) |  |

Note: The Muslim religion could be a risk factor insofar as it supports polygamy. frequenting sex workers increases the risk factors, as does not using protection with a new partner. these are risky behaviours.

## DISCUSSION

The study found an overall HIV/AIDS prevalence of 3.8% (2022), which was statistically different (P<0.001) from the 1.55% reported by DHIS 2018 in these two regions [7]. The incidence rate of 1.63% was also statistically significantly higher (P<0.01) than the 0.24% reported by DHS 2018 in Cameroon. The high prevalence rate may be attributed to the survey being conducted in hospitals. However, the prevalence rate of 2.7% in Cameroon, as estimated by DHS in 2018 [7], is reflected in this national statistic. The study found that the incidence of HIV infection was 1.68%, which is significantly higher than the national incidence in Cameroon (0.24% according to DHS in 2018) with a p-value of less than 0.001. Factors such as religion, engaging in sex work, having unprotected sex with a new partner, and having STIs (syphilis and hepatitis B infections) were statistically significant in explaining this high incidence. These findings are consistent with previous studies conducted among pregnant women or blood donors [13–16]. Recent studies indicate a re-emergence of syphilis [17] (1.02% incidence for an overall prevalence of 1.17%). Syphilis is known to promote the acquisition of HIV through the sexual lesions it causes [18–21].

The incidence of health issues in these regions may be worsened by the inadequate health system and insecurity, which have been intensified by the recent influx of refugees. This has led to increased promiscuity, poverty, and unsanitary conditions. Several studies have focused on the impact of armed conflict and displaced populations, demonstrating the negative effects of political instability on HIV/AIDS response [22]. Although reviews indicate insufficient evidence to conclude that armed conflict is associated with an increase in prevalence [23], our study, although conducted on a smaller population, shows an increase in prevalence in the specific context of Cameroon. In Cameroon, the health coverage falls short of the WHO’s recommended ratio of one health center per 10,000 inhabitants, with only one center per 20,000 inhabitants [11]. This per inhabitant ratio drops again with displaced neighbour populations and this worsen health quality.

Ankouane et al. (2016) conducted a study on blood donors at the Yaoundé Central Hospital and found a prevalence of syphilis to be 0.2% [15]. Newman and colleagues (2015) reported a worldwide prevalence of treponemal infection to be 0.5-0.6% [24]. Recent research suggests that syphilis is re-emerging globally [21,25–28]. Treponemal infection has the potential to act synergistically with HIV, increasing infectivity [18–20,29,30]. The reappearance of syphilis in regions with lower HIV prevalence in Cameroon is a cause for concern, as it suggests the possibility of new HIV infections.

The prevalence and incidence of Hepatitis B were 4.02% and 4.72%, respectively. These data demonstrate a reciprocal correlation between the occurrence of infection with HIV, as seen in prevalence and incidence studies conducted in co-infected individuals [31]. The spread of the disease could be attributed to socio-cultural lifestyle factors such as eating in a common dish, sharing same needle for scarification, as well as reluctance to seek treatment, resulting in an increase in new infection cases caused by the Hepatitis B virus [32]. To prevent the spread of hepatitis B, community education and counseling are necessary. The age group with the highest infection rate was between 25 and 35 years, which corresponds to the most sexually active age group, as supported by other national and international data [2,33,34]. Muslims had a higher infection rate (n=8; P<0.03) compared to Christians. This data supports a study conducted by Mboppi et al. in 2014 in Meyomessala, South Cameroon [8]. The study found that the Islamic religion is more accepting of polygamy and the establishment of a patriarchal society. Nesamoney et al. conducted an epidemiological and observational study in Zambia that suggests religion [35] and ethnicity/race [36], particularly the associated practices, are significant factors in understanding the trend of HIV infection among Muslims. One of the socio-behavioural factors that facilitates HIV acquisition is having had unprotected sex (n=5; P<0.007). Previous incidence and prevalence studies [25,37–39] have shown that factors that aggravate infection or the occurrence of infection are generally related to an individual’s sexual behavior in society. Sex workers are a fraction of the population that is constantly exposed and vulnerable to infection [25,36,40], as well as are people with disabilities. [41] Therefore, individuals who engage in sexual activity with this group are at risk of contracting HIV and other sexually transmitted infections. The study found that men who engaged in sexual activity with female sex workers (n=9) were more likely to contract HIV than those who did not associate with these women (n=5; P<0.0001).

In WHO releases key facts on HIV, **“**HIV continues to be transmitted in every country in the world, with some countries reporting an upward trend in new infections after a period of decline **». https://www.who.int/news-room/fact-sheets/detail/hiv-aids 13 july 2023**. We acknowledge that our study is limited by the size of the population. However, the observed trend is concerning.

Participants were recruited from regional hospitals in the North and Far North, which may introduce bias and potentially inflate the infection rate, as found in the study.

Although the study period of 12 months is relatively short compared to other multi-year impact studies [42,43], the observed trend is significant.

## CONCLUSION

This study shows that HIV infection is resurgent in the North and Far North regions of Cameroon, mainly due to the increasing prevalence of syphilis and hepatitis B infections, in addition to religion and risky sexual behavior. The findings also link increased HIV prevalence to insecurity and population displacement. This trend may be reproduced in the same contexts in other countries where political instability prevails. It is therefore necessary to pay more attention to the control of the epidemic so that the efforts made are consolidated and not washed away.

## Data Availability

Data collected are fully available and presented in the manuscript.

## Acknowledgments

to all participants who have taken part in the one-year study.

## Notes

### Competing Interest Statement

The authors have declared no competing interest.

### Funding Statement

Cameroon Government, through the Chantal Biya International Reference Centre. The funder play no role in the project

### Author Declarations

This work was approved by the hospitals and the National Ethics Committee for Human Health in Cameroon (N°2021/06/85/CE/CNRESH/SP). Recruited participants agreed to participate in the study and signed the informed consent form after reading and understanding it.

